# SonoPatch: Wearable Sonophoresis for On-Demand Physiological Modulation

**DOI:** 10.64898/2026.07.03.26357138

**Authors:** Kye Shimizu, Nathan Whitmore, Yuyang Zhang, Aima Hossen, Pattie Maes

## Abstract

Existing interfaces modulate user experience through visual, auditory, and haptic channels, but direct physiological modulation, which programmatically alters a user’s internal state, remains largely underexplored. We present a wearable sonophoresis patch that uses low-frequency acoustic stimulation to deliver psychoactive substances transdermally, and evaluate its potential for programmable physiological modulation in HCI. We tested this in a double-blinded study (N=26) delivering 100 mg caffeine versus sham control, recording physiological signals during rest and a sustained attention task (SART). The planned comparison for heart rate standard deviation during rest was significant (HR SD *p* = 0.025, *d* = 1.48), with the caffeine group showing suppressed HR SD consistent with sympathetic activation. Mean heart rate at rest was not significant (*p* = 0.365), but exploratory analyses during the cognitive task revealed significant cardiovascular divergence: heart rate (*p* = 0.003) and heart rate standard deviation (*p* = 0.027) both moved in directions consistent with systemic caffeine delivery, with effects emerging within minutes of device activation and a sustained group effect across all task rounds (*p* < 0.001).

These results provide indirect evidence that wearable sonophoresis can deliver substances to modulate user physiology, opening the design space for on-skin chemical interfaces that adapt delivery in real time to change the user’s physiological state on demand.

## 1 Introduction

HCI has progressively expanded the channels through which interactive systems can influence the user. Visual, auditory, and haptic interfaces change what the user perceives; more recent work has moved toward changing the user’s physiological state itself, through electrical muscle stimulation, vestibular stimulation, and autonomic modulation [13, 25, 39]. This trajectory points toward a broader vision: systems that not only sense the body but actively intervene in it.

Chemical HCI extends this vision to the body’s own signaling molecules. Because the vast majority of signaling in the human body is mediated through hormones, neurotransmitters, cytokines, and other chemical messengers, chemical interfaces have the potential to go beyond surface-level sensation and support health, wellness, and cognition by allowing electronic control of physiological processes.

A key application is more precise drug delivery with fewer side effects. A major challenge in conventional pharmacology is delivering drugs exactly when they are needed: most require significant time to be absorbed, and the body’s processing of a substance follows a fixed trajectory that is difficult to control once a dose is taken [55]. The difficulty of controlling drug concentrations can increase the risk of side effects and decrease the effectiveness of a drug. For example, fluctuation in the concentration of anti-Parkinson’s drugs in the body often reduces their effectiveness [43]. In other cases, side effects occur when a drug persists too long in the body; for example many stimulants cause sleep problems because blood levels remain high during the night [33]. Chemical HCI approaches could address the onset problem by triggering delivery precisely when needed, rather than relying on a dose taken minutes or hours in advance.

Ideally, such a system would allow a drug to be introduced rapidly and on demand, be compatible with a wide range of substances, be safe and tolerable for extended use, and be capable of administering a substance without user action. No existing drug delivery system meets all of these criteria in humans. However, sonophoresis, which uses low-frequency acoustic stimulation to temporarily increase skin permeability [37, 45], offers a promising alternative. Unlike oral ingestion, sonophoresis can be toggled programmatically, and because the mechanism acts on the skin itself rather than on any particular molecule, the same hardware could deliver different compounds by changing the solution on the patch [45]. In-vitro and animal studies have demonstrated enhanced transdermal transport of caffeine, insulin, and other molecules [4, 5, 38], but no prior work has tested whether sonophoresis can produce rapid, measurable physiological changes in living humans.

Based on these advantages, we tested whether a wearable sonophoresis device could deliver caffeine transdermally and produce measurable changes in physiology and behavior. Caffeine is an ideal probe because its cardiovascular effects (increased heart rate, reduced heart rate standard deviation) are well characterized and controllable [33, 52], making it possible to infer successful delivery from wearable physiological measurements alone. We validate this in a controlled, double-blind evaluation with 26 participants.

### 1.1 Contributions

In this study, we demonstrate that sonophoresis with a wearable device can be used to rapidly control physiology by delivering a drug through the skin. Through a controlled evaluation with 26 participants, we find that the pre-registered prediction that caffeine sonophoresis would reduce heart rate standard deviation was significant (HR SD *p* = 0.025, *d* = 1.48), consistent with a caffeine-induced autonomic state change detectable by wearable sensors. Exploratory analyses during a cognitive task revealed further cardiovascular divergence (heart rate *p* = 0.003; HR SD *p* = 0.027; mixed model group effect *p* < 0.001).

## 2 Related Work

### 2.1 Transdermal Delivery and Sonophoresis

Transdermal delivery avoids the variability of oral ingestion and sends compounds directly into the bloodstream without the delays of digestion [45]. The challenge is that the stratum corneum (the skin’s outermost barrier layer) is designed to keep molecules out. Existing approaches each solve this differently, with distinct trade-offs for programmability and versatility. Passive patches (nicotine, fentanyl) deliver at a fixed rate with no on-demand control [49]. Microneedle patches physically puncture the skin but are invasive and typically single-use [48]; recent programmable microneedle systems [65] demonstrate digital control but still require skin puncture. Insulin pumps deliver continuously varying doses but require aseptic cannula insertion and are limited to a single pre-loaded drug [42]. Smart capsules [34] are ingested and release their contents under programmed conditions, but offer limited control over timing and dose rate once swallowed.

Sonophoresis is distinctive among these approaches because it is non-invasive, electronically controllable, and substance-agnostic. The mechanism is well characterized: below roughly 20 kHz, acoustic energy creates microscopic bubbles at the skin surface that collapse and temporarily open transport pathways through the skin barrier [36, 58, 59]. The barrier recovers within hours [45]. Because the acoustic energy acts on the skin itself rather than on any particular molecule, the same device can deliver different compounds simply by changing the applied solution. This has been demonstrated in vitro and in animal models with insulin, lidocaine, fentanyl, and caffeine [5, 44, 45, 67]. Relevant to this study, Boucaud et al. [5] showed that low-frequency ultrasound increases caffeine transport across excised human skin, and caffeine reaches the bloodstream intact regardless of delivery route [30].

Despite these promising results, prior sonophoresis research has focused on skin permeability and drug transport rather than downstream physiological effects. No study has tested whether sonophoresis can produce rapid, measurable physiological changes in living humans. Our experiment addresses this gap.

### 2.2 From Body Sensing to Body Actuation in HCI

Fairclough [13] formalized the concept of the “biocybernetic loop”: systems that sense physiological state, infer the user’s condition, and intervene to shift it. As introduced above, a growing line of HCI work has moved from sensing toward actuation, progressing through several levels of intervention depth.

At the shallowest level, systems alter what the user *perceives* without changing underlying physiology: topical chemicals that produce thermal or tingling sensations [28], trigeminal scents [8], AR flavor overlays [41], chemical taste modulators that retarget the flavor of real foods [29], and thermal elements [66]. Brooks et al. [7] surveyed this emerging design space of smell, taste, and temperature interfaces, highlighting how chemical stimuli can serve as a compact, low-power actuation channel. Amores and Maes [1] demonstrated Essence, a wearable olfactory necklace that delivers scent stimuli based on physiological and contextual data to unconsciously influence mood and cognitive performance. Ghandeharioun and Picard [15] showed that subtle visual and auditory breathing cues (BrightBeat) can slow respiration and cultivate calmness without requiring focused attention. Jain et al. [20] formalized this direction with a tripartite model of consciousness for wearable design, arguing that interfaces operating on preconscious and metaconscious pathways can alter emotion, cognition, and behavior without demanding attentional resources. These systems are effective for modulating subjective experience but do not change the body’s internal state.

A deeper class of systems produces genuine physiological state change by stimulating the body’s own effectors. Jain et al. [19] demonstrated that pneumatic stimulation of carotid baroreceptors can modulate heart rate by acting directly on the autonomic nervous system. Azevedo et al. [3] showed that a wrist-worn heartbeat-like vibration measurably reduced skin conductance and self-reported anxiety. Costa et al. [9] closed this into a full biocybernetic loop, sensing negative affect and delivering haptic cues to regulate emotion and cognitive performance. Brooks and Lopes [6] used thermal feedback in the nose to implicitly guide breathing rate without the user’s awareness. Heffernan et al. [16] surveyed the emerging practice of self-implanting NFC and RFID chips under the skin, pointing toward a dissolution of the boundary between body and computer.

All of these systems actuate through physical energy (electrical current, mechanical pressure, vibration, heat). Chemical delivery represents the next step: rather than stimulating a nerve or receptor, the system delivers a molecule that changes the body’s biochemistry directly, with effects that persist after the device stops.

### 2.3 Timing-Dependent Cognitive Enhancement

The broader health behavior literature has formalized the idea that interventions are most effective when delivered at the right moment. Just-in-time adaptive interventions (JITAIs) use real-time sensor data to deliver support precisely when a person is both vulnerable and receptive [40]. JITAIs have been deployed for smoking cessation [17], physical activity [21], and stress management [53], typically delivering digital nudges (notifications, breathing exercises) rather than physiological interventions. Mishra et al. [35] used contextual and behavioral signals to predict when users are receptive to mobile health interventions, showing that sensor-driven timing can improve intervention effectiveness. The limitation shared by the majority of current JITAIs is that the intervention itself is informational or perceptual: the system can tell you to take a break or guide a breathing exercise, but it cannot directly change your biochemistry.

Caffeine’s cognitive benefits illustrate why timing matters for chemical interventions specifically. Effects are highly dependent on when caffeine is consumed relative to the user’s circadian state and fatigue level [33]. Vital-Lopez et al. [63] developed a computational algorithm that optimizes caffeine scheduling based on sleep history, demonstrating that optimized timing alone could improve alertness by 64% or reduce consumption by 65%. Their follow-up 2B-Alert system [62] provides personalized real-time caffeine recommendations via a mobile app, learning individual sleep phenotypes to prescribe both amount and timing. These systems show that the delivery mechanism matters as much as the substance: the right amount of caffeine at the wrong time is wasted, and the right time with the wrong delivery mechanism (15–45 minute oral onset, variable absorption depending on stomach contents) means the window of opportunity may pass before the effect arrives.

Combining these two threads, a chemical JITAI would sense physiological state, decide that intervention is needed, and deliver a substance at the moment it will be most effective. The sensing and decision-making components exist; what is missing is a delivery mechanism that is non-invasive, programmable, and fast enough to act within the window of opportunity.

### 2.4 Positioning Our Work

Prior work provides the key components: body actuation systems that change physiological state through physical energy, sonophoresis research demonstrating enhanced skin permeability in vitro and in animals, and JITAI frameworks showing that timely intervention improves outcomes. However, no study has tested whether sonophoresis can produce rapid, measurable physiological changes in living humans.

Our study addresses this specific gap. We test whether a compact, wirelessly controlled sonophoresis patch can deliver caffeine transdermally at levels that produce cardiovascular changes detectable by consumer wearable sensors. We do not implement closed-loop delivery, test multiple substances, or build a full chemical JITAI. Rather, we provide initial human evidence that the delivery component works, which is a prerequisite for the context-aware, multi-substance systems envisioned above. To our knowledge, no prior system has combined non-invasive transdermal enhancement with wireless programmable control and demonstrated downstream physiological effects in humans.

## 3 Methods

### 3.1 Study Design and Participants

We conducted a double-blind, randomized, placebo-controlled between-subjects study (N=26, n=13 per group) evaluating transdermal caffeine delivery via a wearable sonophoresis device. Sonophoresis uses low-frequency acoustic stimulation to enhance skin permeability for non-invasive substance delivery [32, 37]. The protocol received IRB approval (Protocol #2508001759) after ethics review, clinical research trial review, and environmental health and safety review.

Twenty-six healthy adults were recruited from the local university community (see Section 4 for demographic breakdown). Eligibility required age 18–35 with no caffeine consumption within 3 hours of the session. Exclusion criteria included cardiovascular disease, anxiety disorders, hypertension, tattoos on the dominant forearm, pregnancy, diabetes, and use of antidepressants, anticoagulants, or benzodiazepines. Participants received either 100 mg caffeine (active), roughly equivalent to one 8 oz cup of brewed coffee [61], or 100 mg creatine monohydrate (sham) via sonophoresis. Creatine was selected as the sham substance because (1) both powders are white and odorless and produce visually indistinguishable solutions when dissolved in water, preserving the double-blind; and (2) unlike caffeine, creatine lacks acute sympathomimetic or psychoactive effects at this dose, so it does not confound the primary outcome measures (heart rate, HR SD, EDA, skin temperature) [23]. Both groups received identical acoustic drive from the sonophoresis patch (16 kHz, same duration and power), so any vibration or audible artifact was the same across conditions; only the dissolved substance differed. One experimenter prepared both solutions while a second blindly selected one, ensuring neither participant nor administering experimenter knew the assigned condition. Compensation was a $40 gift card.

### 3.2 Wearable Sonophoresis Device

The sonophoresis device is a custom wearable patch that drives a Duramobi mini speaker [10] at approximately 16 kHz in direct skin contact on the dominant forearm, controlled wirelessly via an ESP32-WROOM-32 microcontroller [12] over Bluetooth Low Energy. We chose 16 kHz because lower frequencies produce larger cavitation bubbles at the skin surface, which collapse more forcefully and create wider transport pathways through the outer skin layer [58, 59]. Below roughly 20 kHz, the energy threshold needed to trigger this cavitation drops sharply, making low frequencies the most efficient range for enhancing skin permeability [45]. Current-limiting circuitry (max 4 mA) and battery-only operation provide safety redundancy. Figure 2 shows the assembled device worn on the wrist and the internal electronics.

Physiological signals were recorded with an EmotiBit biosensor [11] on the non-dominant wrist (heart rate, HR SD, EDA, skin temperature) and a Muse S EEG headband [18] (frontal channels AF7/AF8). All data streams were synchronized and recorded in XDF format using Lab Recorder [56] via the Lab Streaming Layer (LSL) protocol [22]. Each substance solution contained 100 mg powder dissolved in distilled water: USP-grade caffeine (Lab Alley) [24] or NSF-certified creatine monohydrate (Thorne Research) [60].

### 3.3 Procedure

Sessions lasted approximately 70 minutes (Figure 1). After consent, equipment fitting, and a demographic questionnaire, the session proceeded as follows.

**Figure 1:**
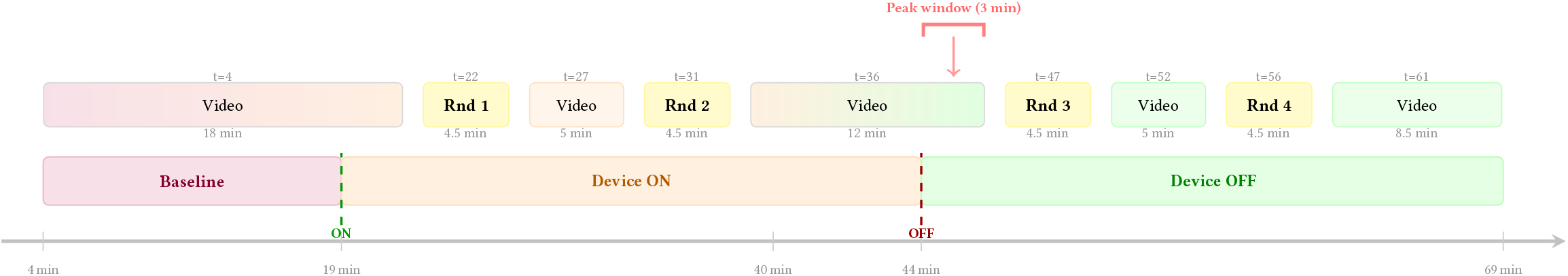
Experimental procedure (training SART round omitted). The time axis shows elapsed time from session start; start times appear above each task box and durations below. Gradient-shaded video segments span phase transitions, indicating the video played continuously while the device was activated or deactivated. The “peak window” indicates the pre-registered measurement window (3-minute pre-Round 3 rest period, starting just after device off). SART rounds lasted ≈4.5 minutes each (225 trials).

**Figure 2:**
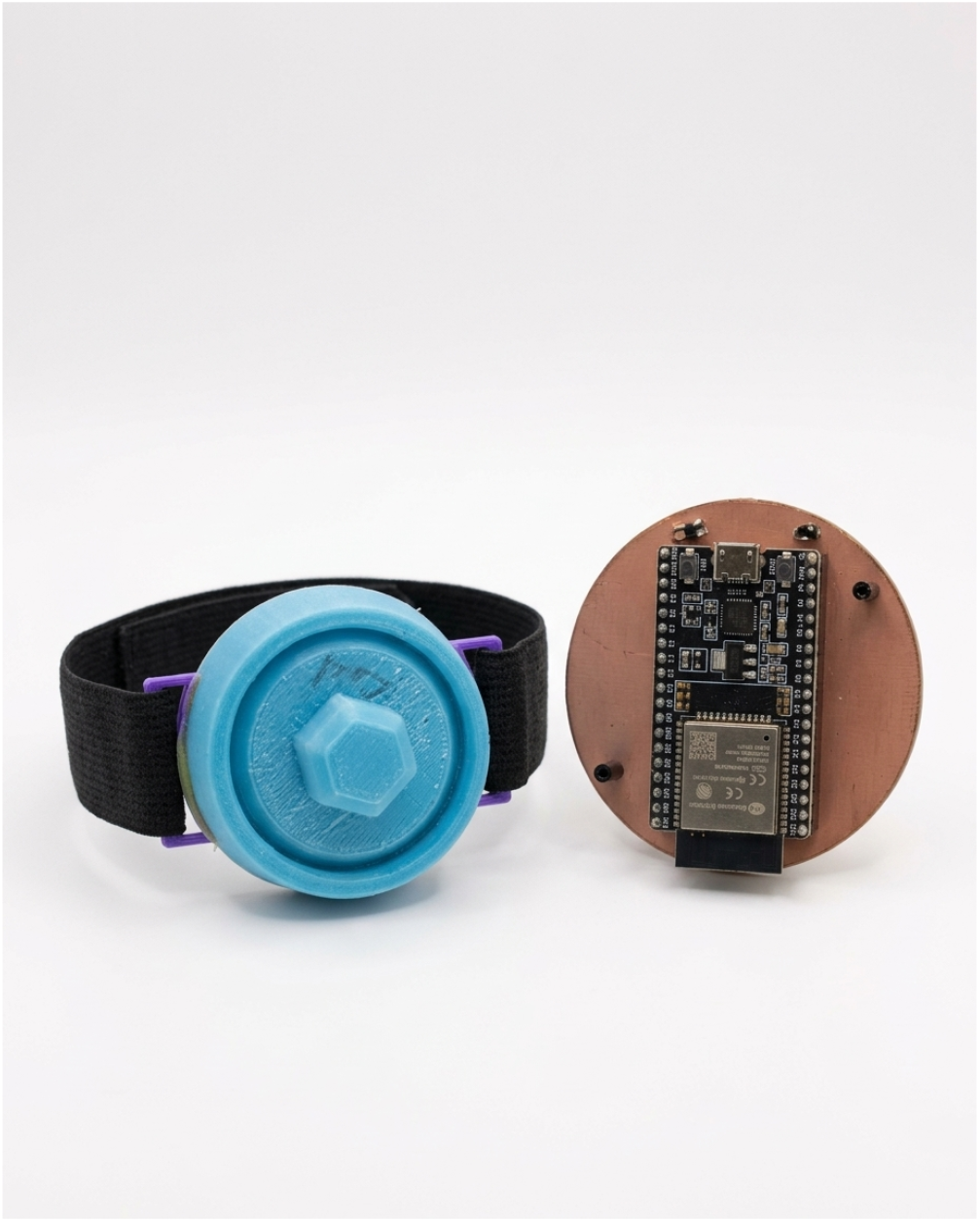
The wearable sonophoresis patch worn on the forearm, with the internal electronics (ESP32 microcontroller on a circular PCB) shown alongside.

**Figure 3:**
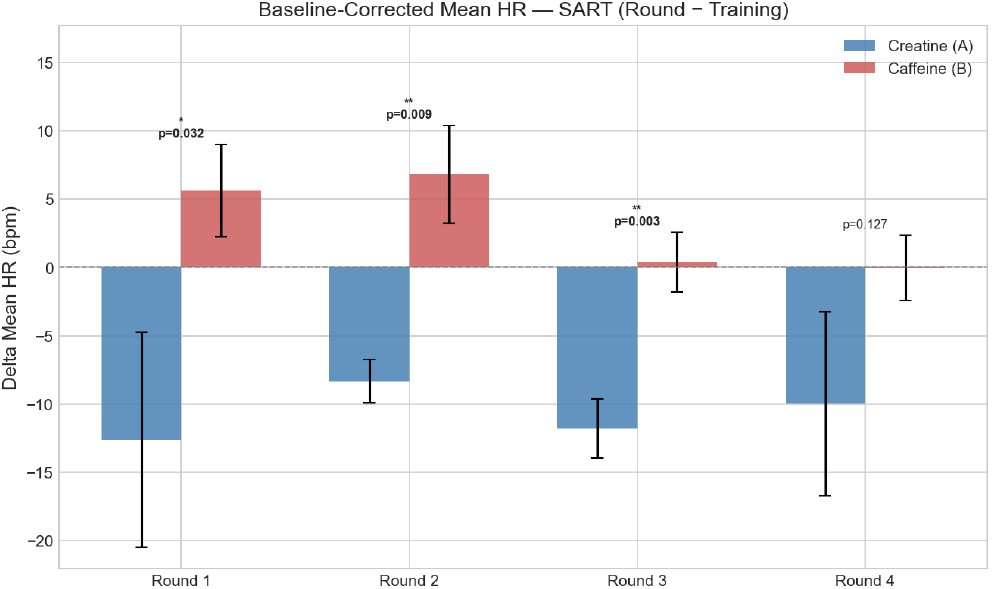
Baseline-corrected mean heart rate during SART task rounds (exploratory analysis). The sham group (blue) declined below baseline across all rounds, consistent with sedentary habituation. The caffeine group (red) maintained heart rate at or above baseline. The group difference was significant at Rounds 1–2 (device ON; *p* = 0.032, 0.009) and Round 3 (device OFF; *p* = 0.003). Error bars show SEM. Note: the pre-registered HR contrast during rest periods was not significant (*p* = 0.365), but HR SD was (*p* = 0.025).

The SART [50, 51] is a go/no-go task in which participants press a button for every digit (1–9) except the target (3), with each digit shown for 250 ms followed by a 900 ms mask (225 trials per round, ≈4.5 minutes each).

#### Training Round (Baseline)

A training SART round established baseline behavioral and physiological performance before substance delivery.

#### Nature Video (15 min)

Participants watched a relaxing nature video to allow physiological signals to stabilize before device activation.

#### Device ON (25 min)

The sonophoresis device was activated. Video segments and SART rounds alternated (3 min video, SART Round 1, 5 min video, SART Round 2, 8.5 min video). Participants may have noticed mild itching or tingling.

#### Device OFF (25 min)

The device was turned off. The same structure continued (3 min video, SART Round 3, 5 min video, SART Round 4, 8.5 min video). Experimenters monitored for adverse reactions throughout and sent a follow-up email 24–48 hours post-session [33, 52]; no adverse events were reported.

### 3.4 Pre-registered hypotheses

In summary, the experiment is a double-blind, between-subjects design in which participants abstained from caffeine for at least 3 hours before the session. After a training SART round and a 15-minute baseline video, the sonophoresis device was activated for 25 minutes (during which participants completed SART Rounds 1– 2, interleaved with video rest periods) and then turned off for a further 25 minutes (Rounds 3–4, same structure). Heart rate, HR SD, EDA, and skin temperature were recorded continuously via the EmotiBit wrist sensor; EEG was recorded via the Muse S headband; and behavioral performance (commission errors, reaction time) was captured during each SART round.

This study was pre-registered prior to analysis [2]. The following hypotheses compared baseline-normalized values at the pre-Round 3 rest period (the 3-minute video-watching window between Round 2 and Round 3, after device deactivation) between caffeine and placebo (creatine) groups:

**H1**. Caffeine will reduce commission error rate (i.e., the proportion of no-go trials where participants incorrectly responded) compared to placebo.

**H2**. Caffeine will increase heart rate compared to placebo.

**H3**. Caffeine will reduce heart rate standard deviation compared to placebo.

**H4**. Caffeine will increase EEG brain rate (spectral centroid) compared to placebo.

Secondary and exploratory hypotheses:

**H5**. Caffeine effects will decrease after the device is turned off.

**H6**. Caffeine effects will interact with habitual caffeine consumption.

Primary outcome measures were mean heart rate and heart rate standard deviation (EmotiBit HR stream, rest periods), commission error rate (SART no-go trials), and the spectral centroid of the frontal EEG from 0.5–30 Hz, which represents an overall measure of alertness and mental arousal [46].

### 3.5 Statistical Analysis

#### Primary contrasts (H1–H4)

For each outcome measure (commission error rate, mean heart rate, heart rate SD, and EEG spectral centroid), a baseline-corrected change score was computed as (pre-Round 3 value −baseline value) for each participant. For physiological measures, the pre-Round 3 rest period is a 3-minute video-watching window between Round 2 and Round 3, and the baseline is the Training SART round. For behavioral measures (commission errors), the baseline is also the Training SART round. Between-group differences were tested using a two-tailed unpaired t-test, or a Wilcoxon rank-sum test when the Shapiro-Wilk test indicated non-normality in either group (*p* < 0.05), as pre-registered.

#### Secondary temporal analysis (H5)

A linear mixed model tested whether the interaction of time point and group predicted changes in physiology or performance across the four baseline-corrected rest-period epochs (pre-Round 2, pre-Round 3, pre-Round 4, post-Round 4). All values were normalized to each participant’s baseline.

#### Exploratory analysis (H6)

The interaction of habitual caffeine consumption and group was tested via linear regression.

#### Exclusions

As pre-registered, participants were excluded if they withdrew or if the stimulator malfunctioned. Three participants whose sonophoresis speaker battery may have failed during the session were excluded from all analyses (physiological and behavioral), as they did not recieve the full dose as per protocol. This yielded 23 participants (11 sham, 12 caffeine) for behavioral analyses. Beyond this, physiological exclusions were applied per modality based on XDF recording quality: an additional 4 participants were excluded from all EmotiBit-based analyses (no EmotiBit data or recording died within 30 minutes), yielding 7 sham and 9 caffeine for EDA and temperature; and 4 more were excluded from heart rate analyses specifically (HR stream with <200 samples), yielding 5 sham and 8 caffeine for heart rate. Only 5 participants (1 sham, 4 caffeine) had sufficient EEG data quality to calculate change in brain rate; descriptive results are reported but no inferential test was conducted due to the insufficient sample. All analyses used *α* = 0.05. No correction for multiple comparisons was applied to the primary contrasts (H1–H4), as each tests a separate pre-registered hypothesis on a different outcome measure. The exploratory temporal analyses across additional time points should be interpreted with appropriate caution, as these were not corrected for the number of windows tested.

## 4 Results

Twenty-six participants completed the study (age *M* = 19.5, *SD* = 1.3; 13 per group). The groups did not differ significantly in age, gender distribution, or habitual caffeine consumption (full demographics in Appendix .1).

### 4.1 Cardiovascular Measures (Primary Outcomes)

#### 4.1.1 Planned Comparisons (H2, H3)

The planned primary analysis specified baseline-corrected change scores during the pre-Round 3 rest period (3-minute video-watching window between Round 2 and Round 3) for mean heart rate (H2) and HR SD (H3), comparing sham and caffeine groups.

For mean heart rate (H2): sham Δ = −6.2 bpm vs. caffeine Δ = + 0.3 bpm (*t* -test, *p* = 0.365); the planned comparison was not significant. For HR SD (H3): sham Δ = + 5.7 bpm vs. caffeine Δ = −6.2 bpm (*t* -test, *p* = 0.025, *d* = + 1.48); the caffeine group maintained lower heart rate standard deviation relative to sham, consistent with the predicted sympathetic activation effect.

#### 4.1.2 Exploratory Analysis: Cardiovascular Effects During Cognitive Load

Cardiovascular measures also diverged significantly between groups during the cognitively demanding SART task. Rounds 1–2 were performed while the device was ON (active delivery); Rounds 3– 4 were performed after the device was turned OFF. After baseline correction (subtracting each participant’s Training value), the sham group exhibited a natural heart rate decline of 8.3 to 12.6 bpm below baseline across Rounds 1–4, while the caffeine group maintained heart rate at or above baseline (−0.04 to +6.8 bpm).

- Round 1 (device ON): sham Δ = −12.6 bpm vs. caffeine Δ = +5.6 bpm (*t* -test, *p* = 0.032)
- Round 2 (device ON): sham Δ = −8.3 bpm vs. caffeine Δ = +6.8 bpm (*t* -test, *p* = 0.009)
- Round 3 (device OFF): sham Δ = −11.8 bpm vs. caffeine Δ = +0.4 bpm (*t* -test, *p* = 0.003)
- Round 4 (device OFF): sham Δ = −10.0 bpm vs. caffeine Δ = −0.04 bpm (*t* -test, *p* = 0.127)

HR SD during SART task windows also showed a significant group difference at Round 3: sham Δ = + 4.0 bpm vs. caffeine Δ = −3.8 bpm (*t* -test, *p* = 0.027). The caffeine group’s heart rate standard deviation was suppressed relative to sham, a pattern consistent with sympathetic activation.

These task-window effects were not pre-registered (the pre-registration specified rest-period video epochs), but the pattern is notable: the cardiovascular signature of caffeine emerged under cognitive load, when sympathetic drive is elevated, rather than during passive rest. The mixed model confirmed a sustained group difference across all four SART rounds (group effect: +18.1 bpm, *p* < 0.001; interaction *p* = 0.229).

#### 4.1.3 Temporal Profile (H5)

We fit linear mixed models (group × time, random intercept per participant) to test whether effects changed over time. For mean HR during SART rounds, the group effect was significant (+ 18.1 bpm caffeine vs. sham, *p* < 0.001) with no group × time interaction (*p* = 0.229), indicating a consistent difference across rounds (Figure 4). HR SD during SART rounds showed no significant effects (group *p* = 0.117; interaction *p* = 0.909). HR SD during rest-period epochs showed no significant group effect (−3.9 bpm, *p* = 0.330) or group × time interaction (*p* = 0.948).

**Figure 4:**
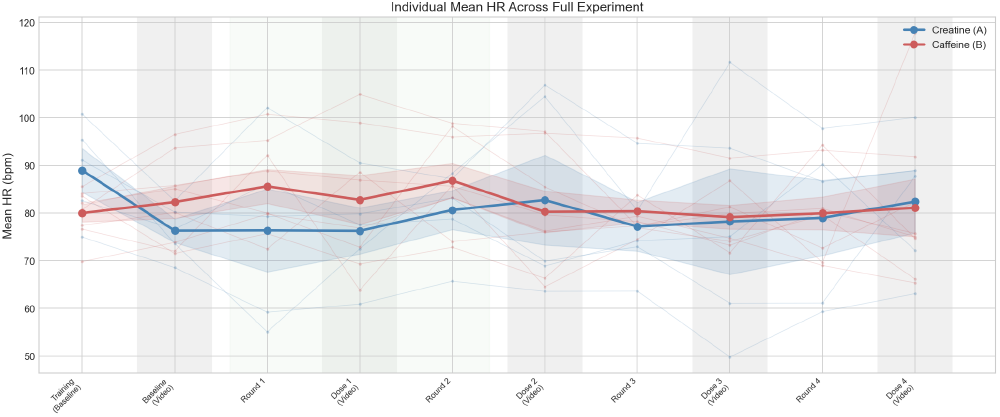
Individual and group-mean heart rate trajectories across the full experiment. The caffeine group (red) maintained heart rate near baseline, while the sham group (blue) declined progressively. Thin lines show individual participants; thick lines show group means with 95% confidence bands. Shaded columns indicate SART task windows; unshaded regions are rest-period video epochs.

#### 4.1.4 Moderation by Habitual Caffeine Consumption (H6)

We used a linear regression to test whether the interaction of group and habitual caffeine consumption (cups/day) predicted cardiovascular measures at the pre-Round 3 rest period. Neither the group × cups interaction for HR (*β* = 3.84, *p* = 0.326) nor for HR SD (*β* = 1.33, *p* = 0.402) reached significance at the pre-registered window.

In exploratory analyses during the SART task (Round 3), the regression model for HR SD was significant overall (*F* (3, 9) = 5.83, *p* = 0.017, *R*^2^ = 0.66), with a significant main effect of group (*β* = −10.3, *p* = 0.008) and cups (*β* = −2.2, *p* = 0.032), though the interaction term remained non-significant (*β* = 1.2, *p* = 0.387). Because the group × cups interaction was not significant, we cannot conclude that habitual caffeine consumption moderated the effect of the patch. However, habitual consumption was independently associated with greater HR SD suppression during cognitive load, regardless of group.

To explore this further, we performed a median split on habitual consumption (high ≥1 cup/day, *n* = 10; low = 0 cups/day, *n* = 13). Within the caffeine patch group, high habitual consumers showed a trend toward greater heart rate elevation at Round 2 (+ 14.9 bpm vs. +2.0 bpm; Figure 5) and greater HR SD suppression at Rounds 3–4 (Figure 6). This pattern did not appear in the creatine group, suggesting that habitual intake may interact with transdermal caffeine delivery, though the formal interaction test did not reach significance.

**Figure 5:**
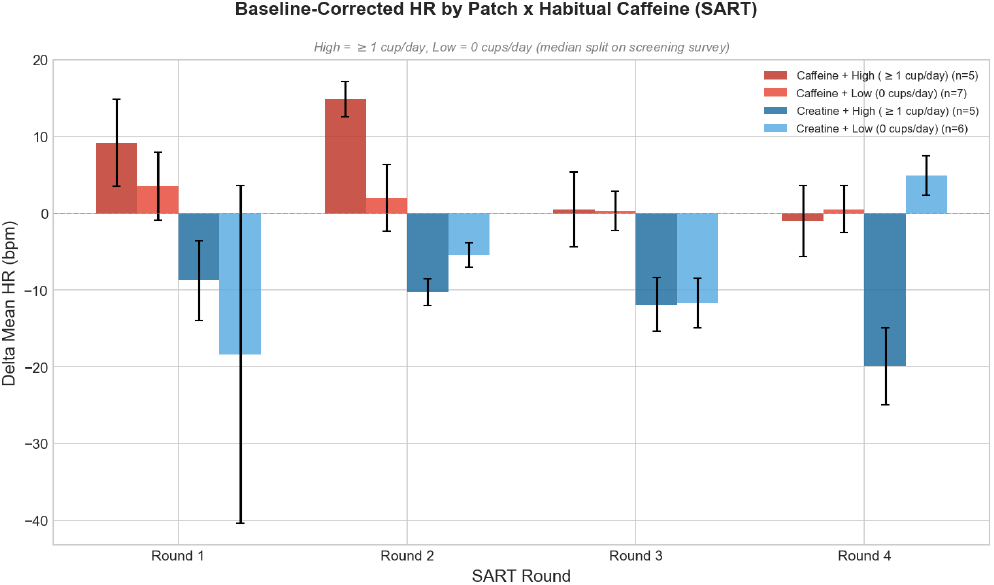
Baseline-corrected mean heart rate by patch group and habitual caffeine consumption (median split: high ≥1 cup/day, low = 0 cups/day). Within the caffeine group, high habitual consumers (dark red) showed a trend toward greater HR elevation at Round 2 (*p* = 0.076). No moderation appeared in the creatine group.

**Figure 6:**
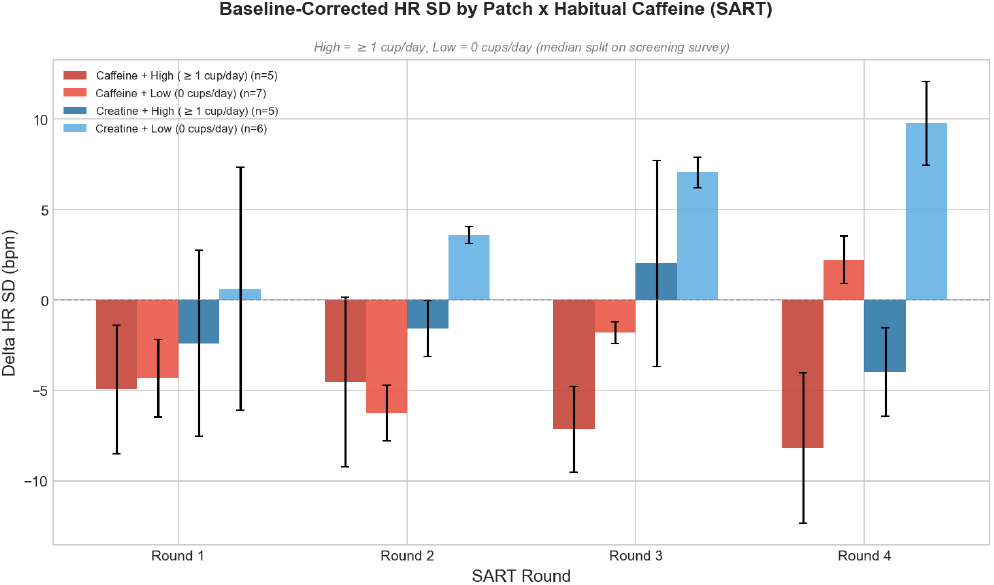
Baseline-corrected HR SD by patch group and habitual caffeine consumption (median split: high ≥1 cup/day, low = 0 cups/day). Within the caffeine group, high habitual consumers (dark red) showed significantly greater HR SD suppression than low consumers (light red) at Round 3 (*p* = 0.031) and Round 4 (*p* = 0.025). At Round 2, both subgroups showed similar suppression. This pattern did not appear in the creatine group.

### 4.2 Electrodermal and Temperature Measures

EDA and temperature analyses included 7 sham and 9 caffeine participants after per-modality exclusion. Tonic EDA and phasic EDA showed no significant group differences at any time point (all *p* > 0.10). Skin temperature during rest periods moved in opposite directions for the two groups: the sham group cooled from baseline (Δ = −0.09 ^°^C at pre-Round 3) while the caffeine group warmed (Δ = + 0.27 ^°^C), though the difference was not significant (*p* = 0.295). This divergence is consistent with caffeine’s known thermogenic effect at rest [33], though the sample was too small to reach significance.

### 4.3 EEG Measures

After rejecting bad channels and data points we computed the change in spectral centroid of the frontal EEG (channel AF7 or AF8; the channel was selected for each participant and time window based on the channel with the highest signal.). Rejection was performed as per the preregistration; first the optimal channel was selected (or the participant was excluded if neither channel contained good data), then the data were epoched to encompass rest periods and time-domain artifact rejection (EEGlab ASR) was used to reject periods of time containing artifacts. Finally, we calculated the spectral centroid of the artifact-free data for the training period and the pre-Round 3 video rest period and calculated the change for each participant.

Only 5 participants (and only 1 control participant) had sufficient EEG data quality in both the training and pre-Round 3 windows to permit analysis. The control participant had an increase of 2.36 Hz in their spectral centroid frequency, while caffeine participants universally showed a decrease (mean -1.65 Hz, SD=1.29). As there was only one control participant with data, we could not conduct a statistical analysis as originally planned.

### 4.4 Behavioral Measures (H1)

SART performance did not differ between groups (Table 2). Accuracy was near ceiling in both conditions (≈95%), and reaction times showed no differential change over time. The sham group had higher baseline commission error rates (*M* = 59.3%) than the caffeine group (*M* = 40.4%, *p* = 0.065), but this difference was present at Training before device activation. After baseline correction (subtracting each participant’s Training commission error rate), no significant group differences emerged at any round (all *p* > 0.14; Figure 7). The sham group showed a practice-related decline in commission errors across rounds, while the caffeine group remained stable. Full per-round statistics are in Table 2 (Appendix .2).

**Table 1:** Pre-registered hypotheses and outcomes. Planned comparisons used the pre-Round 3 rest period (3-min video window between Round 2 and Round 3). ^*^Not significant at the planned rest-period window, but significant during the exploratory SART task analysis.

| H | Measure | Result (rest; SART) | Sup. | Sec. |
| --- | --- | --- | --- | --- |
| H1 | Commission errors | All $p > 0.14$ (baseline-corrected) | No | 4.4 |
| H2 | Heart rate | $p = 0.365$; $p = 0.003$ | No* | 4.1.1 |
| H3 | HR SD | $p = 0.025$; $p = 0.027$ | Yes | 4.1.1 |
| H4 | EEG spectral centroid | Insufficient sample ($n = 5$) | N/A | 4.1.1 |
| H5 | Temporal profile | HR $p = 0.229$; HR SD $p = 0.948$ (group × time) | No | 4.1.3 |
| H6 | Habitual caffeine | $p = 0.402$ (group × cups) | No | 4.1.4 |

**Table 2:** Behavioral measures by group and SART round. Values are mean (SD). Creatine = sham (n=11), Caffeine = active (n=12). Three participants with speaker battery failures excluded. *p*-values from independent *t* -tests or Mann-Whitney *U* .

| Measure | Round | Creatine | Caffeine | $p$ |
| --- | --- | --- | --- | --- |
| Accuracy (%) | Training | 90.6 (7.5) | 92.7 (5.7) | 0.498 |
|  | Round 1 | 94.5 (2.7) | 95.7 (2.2) | 0.246 |
|  | Round 2 | 93.8 (3.9) | 95.7 (2.5) | 0.253 |
|  | Round 3 | 93.9 (4.0) | 94.9 (2.6) | 0.901 |
|  | Round 4 | 94.7 (3.2) | 94.7 (2.9) | 0.640 |
| Commission errors (%) | Training | 59.3 (27.2) | 40.4 (26.4) | 0.065 |
|  | Round 1 | 50.4 (23.1) | 41.5 (21.7) | 0.352 |
|  | Round 2 | 48.0 (19.5) | 40.8 (21.9) | 0.415 |
|  | Round 3 | 48.3 (23.9) | 43.2 (21.9) | 0.593 |
|  | Round 4 | 44.9 (27.9) | 43.1 (22.8) | 0.689 |
| RT mean (ms) | Training | 362.6 (66.7) | 432.5 (61.4) | 0.016 |
|  | Round 1 | 346.2 (45.4) | 385.2 (51.4) | 0.068 |
|  | Round 2 | 344.7 (31.3) | 391.0 (63.9) | 0.103 |
|  | Round 3 | 347.5 (45.3) | 391.3 (78.1) | 0.559 |
|  | Round 4 | 351.1 (41.2) | 399.6 (94.0) | 0.166 |
| RT SD (ms) | Training | 95.2 (25.7) | 105.5 (24.9) | 0.341 |
|  | Round 1 | 91.9 (22.0) | 81.4 (12.2) | 0.166 |
|  | Round 2 | 82.5 (16.5) | 83.5 (16.6) | 0.884 |
|  | Round 3 | 84.0 (23.2) | 86.6 (18.3) | 0.767 |
|  | Round 4 | 82.8 (28.7) | 92.8 (20.2) | 0.132 |

**Figure 7:**
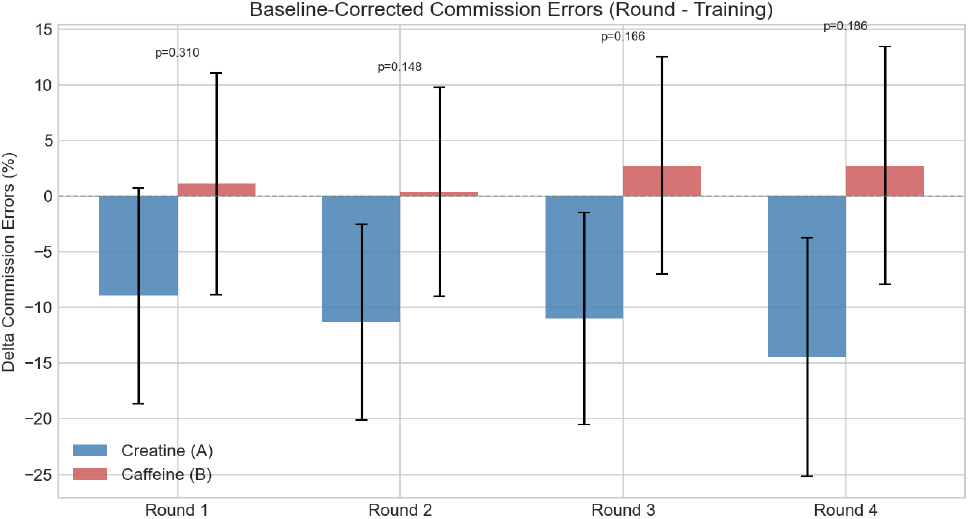
Baseline-corrected commission error rates (Round−Training). The sham group (blue) showed a practice-related decline from their high baseline, while the caffeine group (red) remained near zero. No significant group differences at any round (all *p* > 0.14).

### 4.5 Summary Against Pre-Registered Hypotheses

Table 1 summarizes the outcome of each pre-registered hypothesis. H3 (HR SD) was supported at the planned comparison window: the caffeine group showed significantly lower HR SD than sham during the pre-Round 3 rest period (*p* = 0.025, *d* = + 1.48), and this effect was also significant during the SART task (*p* = 0.027). H2 (HR) was not significant at the planned rest-period window (*p* = 0.365), but exploratory analyses during the SART cognitive task revealed significant group divergence (HR *p* = 0.003 at Round 3), suggesting the mean heart rate effect is most detectable under cognitive load. H5 showed no significant group × time interaction for either measure. H6 showed no significant group × cups interaction at the planned window (*p* = 0.402), but habitual caffeine consumption was a significant predictor of HR SD during the SART task (*p* = 0.032). H1 was not supported: behavioral performance did not differ between groups.

## 5 Discussion

### 5.1 Interpreting the results

The HR SD effect at rest (*p* = 0.025, *d* = 1.48) and the cardiovascular divergence during cognitive load (*p* = 0.003 at Round 3) together indicate that the patch shifted autonomic balance in the caffeine group.^1^ The effect was amplified under cognitive load, when the body’s arousal systems were already engaged, and persisted after the device was turned off, consistent with caffeine’s long duration of action [33]. The absence of a behavioral effect is consistent with prior work showing that cardiovascular effects of caffeine appear at lower doses than cognitive effects [33], suggesting the delivered amount was sufficient to shift heart rate but not enough to affect attention in well-rested participants performing a near-ceiling task. Without direct measurement of caffeine in the body, we cannot confirm the exact absorption timeline; sonophoresis may accelerate onset compared to passive transdermal delivery [30], but this remains to be validated. The descriptive trends in habitual consumers (Section 4.1.4) suggest that identical delivery produces different responses depending on the user’s caffeine history, an important consideration for system design.

#### Time course of effects

We predicted that caffeine sonophoresis would allow for us to rapidly toggle the effects of caffeine on and off because it would bypass the lag associated with gastrointestinal absorption of caffeine (typically reaching peak plasma concentration in 30–60 minutes and producing noticeable behavioral and cardiovascular effects within 15–45 minutes [14, 33]). We also predicted that caffeine effects would diminish rapidly when the sonophoresis was turned off, stopping the flow of caffeine into the body and causing blood levels to begin dropping due to liver metabolism. These predictions were partially confirmed as we observed a rapid onset of caffeine effects within 7.5 minutes of beginning administration (round 1). Importantly, this is much faster than other transdermal caffeine delivery methods such as a patch with no ultrasound, microneedles, or other permeation enhancement; these methods have lag times of hours [30], substantially longer than the timeline observed here.

However, the offset time is less clear. We tested for a significant drop in caffeine effects after turning off the device by testing the interaction of group * time point; we did not observe a significant interaction meaning that caffeine effects did not change significantly over time. Nonetheless, the effect of caffeine on cardiovascular measures converged towards zero at the final SART test (Figure 3), suggesting a possible waning of effects.

These results suggest that the system can introduce drugs rapidly, though the time course of drug effects after the system is turned off remain unclear and likely depend on individual factors like speed of metabolism. Future research could explore the time course using blood chemical sampling to define properties like the half life of caffeine introduced through sonophoresis.

### 5.2 Sonophoresis as an HCI output channel

#### Chemical output vs. physical output

Existing interface modalities (visual, auditory, haptic) change what the user perceives. Drug delivery changes what the user *is*: their autonomic balance, their heart rate, potentially their alertness. This is a different kind of output, one that the user does not perceive during delivery and that gradually declines after the device stops. Prior work has used electrical muscle stimulation to move limbs [26, 27], galvanic vestibular stimulation to alter balance perception [54], and pneumatic baroreceptor stimulation to shift autonomic tone [19]. Chemical delivery extends this trajectory: rather than stimulating a nerve, the system delivers a molecule that changes the body’s biochemistry directly

#### Substance-agnostic delivery

Because the acoustic stimulation disrupts the skin barrier independent of the molecule [37], the same hardware could deliver melatonin, lidocaine, nicotine, glucose, or other compounds by changing the solution. A future device could go further: a multi-reservoir design with separate pouches for different substances, or an integrated microfluidic channel that mixes and routes compounds at programmable concentrations, would allow a single on-body patch to select among substances and adjust their concentration in real time without user intervention. This substance-agnostic property opens a design space for context-aware delivery, where the system selects a substance and dose based on the user’s current physiology and the situational context in which a particular state is desired (e.g., sustained attention, relaxation, pain relief): a productivity system could deliver timed caffeine micro-doses when wearable sensors detect declining attention during focused work; a sleep system could initiate melatonin delivery based on circadian phase and taper it automatically; a pain management system could trigger lidocaine delivery when movement patterns suggest discomfort; and an athletic system could deliver glucose or electrolytes based on exertion level. In each case, the same patch hardware serves as a general-purpose output channel, with the “content” of the output determined by the loaded solution. Because delivery is electronically controlled, the system can vary the quantity and timing of delivery: a short burst for mild alertness, a sustained session for deeper effect, or spaced micro-doses throughout the day, giving software fine-grained control over dosing that passive patches and oral administration cannot provide. Note, however, that this control applies to onset and dose rate; once a substance enters the bloodstream, the speed at which effects diminish depends on the body’s own clearance rate.

#### Bypassing digestion

Transdermal delivery also sends substances directly into the bloodstream, avoiding the delays and variability of oral ingestion. For caffeine this makes little practical difference, since oral caffeine is already absorbed efficiently [30]. But for other compounds, bypassing digestion could increase the effective dose or reduce onset latency. Some substances require processing by the liver to become active, so future applications of this platform with other compounds will need to account for these differences.

### 5.3 Design implications for chemical interfaces

Our results point to several design considerations for systems that deliver substances through the skin.

#### Sensing-actuation coupling

The same off-the-shelf wearable sensors used to monitor participants (EmotiBit for heart rate, EDA, temperature) were able to detect the effect of the delivered substance, establishing a basic sensing-actuation loop. This suggests that a closed-loop system, one that senses physiological state and adjusts delivery accordingly, is feasible with existing consumer hardware. A key design question is latency: transdermal onset is slower than oral ingestion, so the system must predict when a substance will be needed rather than reacting after the fact, shifting the algorithmic challenge from detection to anticipation.

#### Personalization as a first-class requirement

The habitual consumption data show that identical delivery produces different physiological responses depending on the user’s history (Section 4.1.4). This implies that chemical interfaces need user profiling (caffeine history, baseline physiology) or learned dosing models that adapt over time. This parallels adaptive UI research, but with higher stakes: once a substance is delivered, the effect cannot be retracted. A conservative approach would start with low delivery rates and titrate upward based on sensor feedback.

#### Form factor and body placement

Our prototype uses an ESP32 microcontroller and an off-the-shelf mini speaker, but the active components required for sonophoresis (a piezoelectric transducer, a low-power driver circuit, and a small battery) could be miniaturized to a coin-cell-sized adhesive patch. The choice of form factor involves tradeoffs. A watch-style device on the wrist allows easy removal, recharging, and interaction, but limits the skin contact area available for delivery. An adhesive patch on the upper arm or torso offers a larger permeation surface and more consistent skin contact during movement, but is less convenient to apply, remove, and maintain. Different body sites also offer different permeability (behind the ear, inner wrist, upper arm), and designers must balance delivery efficiency against social acceptability, comfort, and whether the device is intended for continuous wear or session-based use. Our forearm placement was chosen for accessibility and consistent contact in a lab setting, not as a recommendation for deployment.

#### Skin coupling and prolonged wear

A practical constraint is that sonophoresis requires a liquid coupling medium between the transducer and the skin to transmit acoustic energy. In our study, the substance was dissolved in distilled water applied directly to the skin surface. Prolonged wet skin contact risks maceration and irritation, which would limit continuous wear to short sessions. Future designs could mitigate this by using sealed hydrogel reservoirs that maintain coupling without exposing the surrounding skin, or by operating in intermittent duty cycles (e.g., 5 minutes on, 15 minutes off) that allow the skin to dry between delivery windows.

#### Fashion integration

Beyond clinical or utilitarian form factors, chemical delivery hardware could be embedded into everyday fashion. A thin-film transducer laminated into a watch band, a ring, or an adhesive patch worn under clothing would be visually indistinguishable from conventional accessories. Jewelry and textile-integrated electronics are already commercially available (e.g., the Oura ring, Project Jacquard’s conductive yarn woven into everyday garments [47]); adding a miniaturized sonophoresis module with a replaceable substance cartridge would extend these platforms from sensing to actuation. Fashion integration also shifts the social framing: a bracelet that delivers melatonin before sleep or caffeine before a meeting is closer to perfume or cosmetics than to medical devices, potentially lowering adoption barriers. Designers could treat the substance cartridge as a consumable element, similar to fragrance refills, letting users select and swap delivery profiles to match their daily needs.

#### User control and feedback

Participants in our study received no perceptible feedback during delivery, which is desirable for non-disruptive use but raises interaction design questions. How does the user initiate, pause, or override delivery: through a phone app, a gesture on the patch, voice, or fully autonomous operation? How does the system communicate that delivery is active or that a target physiological state has been reached, given that the delivery itself is imperceptible? Options include LED indicators, subtle haptic confirmation, or app notifications, each with different tradeoffs for attention and social context. Safety also requires explicit UX: hard limits on cumulative delivery, lockout periods between doses, and clear override mechanisms.

### 5.4 Limitations

Several limitations should be considered when interpreting these findings.

#### Sample Size and Equipment Attrition

While 26 participants completed the study, equipment failures (stimulator battery depletion and recording loss) reduced the physiological analysis sample to 5–10 participants per group depending on the metric. This limited our ability to detect changes in some metrics, particularly the EEG metrics. Despite the small sample, the primary cardiovascular measures reached significance (*p* < 0.05), suggesting robust effects. Future studies should ensure equipment reliability and include larger samples with formal power analyses.

#### Population Characteristics and Individual Variability

Participants were young adults (18–35 years) recruited from a university community, limiting generalizability to older populations or those with different skin characteristics. Skin thickness, hydration, and permeability vary with age and demographic factors, potentially affecting sonophoresis efficacy. Additionally, habitual caffeine consumers may exhibit tolerance effects that differ from caffeinenaive individuals. The between-subjects design also revealed pre-existing baseline differences in some behavioral measures (e.g., commission error rates, reaction time), which may have masked treatment effects. A within-subjects crossover design would control for these individual differences, though it would introduce potential carry-over effects with caffeine.

#### Absence of Direct Delivery Validation

We did not perform *in vitro* permeation testing or measure plasma/saliva caffeine concentrations in this study. However, extensive prior work has validated that that low-frequency sonophoresis can drive transdermal transport of caffeine and other molecules [5, 36, 45]; our results represent an extension of this work showing that caffeine sonophoresis can also drive physiological responses. Future closed-loop systems could incorporate feedback from sweat sensors [57] to measure and respond to changes in the systemic concentration of a substance.

#### Ecological Validity

Laboratory conditions may not reflect real-world usage scenarios where movement, environmental factors, and varied skin conditions could affect device performance and user experience.

### 5.5 Future work

Blood sampling would quantify how much substance reaches the bloodstream and clarify whether the behavioral null reflects an insufficient amount or suboptimal test conditions. A within-subjects crossover design would reduce noise from individual differences but requires sessions separated by at least 24 hours for caffeine effects to dissipate, with careful counterbalancing to preserve blinding.

The closed-loop vision, where sensors detect a need, the system delivers a substance, and sensors verify the effect, is the natural next step. Three problems remain before this is practical: personalization (learning individual response profiles), anticipatory timing (predicting need before it arrives, since transdermal onset is slower than oral), and safety constraints (cumulative limits, inter-delivery intervals, substance interactions). Delegating delivery decisions to autonomous software agents [31, 64] is a natural extension, where an agent manages multiple substances across the day based on physiological context, the same way agents already manage notifications and environmental controls, except the output is chemical rather than digital. Real-world deployment also raises questions about skin variability, sweat, movement, and maintaining consistent contact during daily activities.

## 6 Conclusion

This paper presented a wearable sonophoresis patch and evaluated its ability to deliver caffeine transdermally in a double-blind, placebo-controlled study with 26 participants. The planned comparison for heart rate standard deviation during rest was significant (HR SD *p* = 0.025, *d* = 1.48), with the caffeine group showing suppressed HR SD consistent with sympathetic activation. Exploratory analyses during the SART cognitive task revealed further cardiovascular divergence: the caffeine group maintained heart rate at or above baseline while the sham group declined (HR *p* = 0.003 at Round 3; HR SD *p* = 0.027), with a consistent group effect across all four rounds (mixed model *p* < 0.001). The direction of these effects matches the known autonomic profile of systemic caffeine, and the stronger effects under cognitive load suggest that sympathetic drive amplified the signal.

Behavioral performance on the sustained attention task did not differ between groups, likely reflecting a transdermal concentration below the threshold for cognitive effects in well-rested participants. However, habitual caffeine consumers in the caffeine group showed significantly greater HR SD suppression than non-consumers (*p* = 0.025), consistent with the withdrawal-reversal model discussed in Section 4.

These findings have three implications. **First**, a compact, wirelessly controlled patch can deliver a substance transdermally at levels detectable by off-the-shelf wearable sensors, establishing the basic sensing-actuation loop required for closed-loop physiological systems. **Second**, individual differences in caffeine history and genetics are large enough that any real-world deployment will require personalized dosing, whether through user profiling, learned baselines, or real-time physiological feedback. **Third**, because sonophoresis enhances skin permeability independent of the substance being delivered, the same device architecture could support melatonin, glucose, nicotine, lidocaine, or other compounds simply by changing the solution applied to the skin. This positions the technology not as a single-purpose caffeine delivery system but as a general-purpose transdermal platform, and opens a path toward closed-loop chemical interfaces that monitor a user’s physiological state and deliver the appropriate substance in response.

## Data Availability

All data produced in the present study are available upon reasonable request to the authors

## .1 Participant demographics

Twenty-six participants completed the study (13 female, 12 male, 1 preferred not to say; age *M* = 19.5, *SD* = 1.3, range 18–24). Half reported no daily caffeine consumption (0 cups/day), while the other half consumed 1–7 cups/day (*M* = 1.3, *SD* = 2.0). Participants were assigned to the caffeine group (*n* = 13; 7 female, 5 male; age *M* = 19.7, *SD* = 1.6; caffeine intake *M* = 1.5 cups/day) or the creatine sham group (*n* = 13; 6 female, 7 male; age *M* = 19.4, *SD* = 0.9; caffeine intake *M* = 1.1 cups/day). The groups did not differ significantly in age, gender distribution, or habitual caffeine consumption.

## .2 Behavioral measures

### .2.1 Task accuracy

Overall SART accuracy was high in both groups, with performance near ceiling across all rounds (caffeine: *M* ≈95%; sham: *M* ≈94%). Neither group showed significant changes from training to subsequent rounds, and no significant between-group differences were observed at any time point (all *p* > 0.24).

### .2.2 Commission error rate

Commission errors on no-go trials (digit 3) showed a trend-level baseline difference. The sham group had higher baseline commission error rates (*M* = 59.3%, *SD* = 27.2%) compared to the caffeine group (*M* = 40.4%, *SD* = 26.4%; *p* = 0.065). This difference was present at training before device activation, indicating a pre-existing group difference rather than an experimental effect. The sham group showed a gradual decline in commission errors across rounds (from 59.3% to 44.9%), suggesting a practice effect, while the caffeine group remained stable (≈40–43%). Baseline-corrected comparisons revealed no significant differences between groups at any round (all *p* > 0.14).

### .2.3 Reaction time

Mean reaction time on correct go trials was consistently faster in the sham group (*M* ≈ 350 ms) than the caffeine group (*M* ≈ 400 ms). This difference was significant at Training (*p* = 0.016) but not at subsequent rounds (all *p* > 0.06), and did not change differentially over time, indicating a pre-existing group difference. Reaction time variability (RT SD) showed no significant group differences at any time point (all *p* > 0.13). Omission errors were negligible after training (<0.5% in both groups).

## .3 Physiological statistics

All tables below report baseline-corrected values (value at time point − value at baseline). For physiological measures, baseline is the last 5 minutes of the nature video; for behavioral measures, baseline is the Training SART round. Sham = creatine (Patch A); Caffeine = Patch B. Three participants were excluded from all analyses due to speaker battery failure (see Section 3.5 for the full exclusion criteria). Heart rate analyses include *n*_sham_ = 5, *n*_caff_ = 8 after additional per-modality exclusions (no EmotiBit data or insufficient HR stream); EDA and temperature analyses include *n*_sham_ = 7, *n*_caff_ = 9. Tests used Welch’s *t* -test or Mann-Whitney *U* (MW) depending on Shapiro-Wilk normality; *d* = Cohen’s *d*. ^*^*p* < .05; ^**^*p* < .01; ^†^*p* < .10.

**Table 3:** Baseline-corrected mean heart rate (bpm) during SART task windows. *n*_sham_ = 5, *n*_caff_ = 8.

| Round | Sham $\Delta$ | Caff $\Delta$ | Test | $p$ | $d$ |
| --- | --- | --- | --- | --- | --- |
| Round 1 | $-12.6 \pm 17.6$ | $+5.6 \pm 9.6$ | $t$ | .032* | -1.39 |
| Round 2 | $-8.3 \pm 3.5$ | $+6.8 \pm 10.1$ | $t$ | .009** | -1.82 |
| Round 3 | $-11.8 \pm 4.8$ | $+0.4 \pm 6.2$ | $t$ | .003** | -2.11 |
| Round 4 | $-10.0 \pm 15.1$ | $-0.04 \pm 6.8$ | $t$ | .127 | -0.94 |

### .3.1 Heart rate:SART windows (Round − Training)

**Table 4:** Baseline-corrected HR standard deviation (bpm) during SART task windows. *n*_sham_ = 5, *n*_caff_ = 8.

| Round | Sham $\Delta$ | Caff $\Delta$ | Test | $p$ | $d$ |
| --- | --- | --- | --- | --- | --- |
| Round 1 | $-1.2 \pm 8.0$ | $-4.5 \pm 4.9$ | $t$ | .364 | +0.54 |
| Round 2 | $+0.5 \pm 3.4$ | $-5.6 \pm 5.1$ | $t$ | .040* | +1.33 |
| Round 3 | $+4.0 \pm 7.5$ | $-3.8 \pm 3.7$ | $t$ | .027* | +1.45 |
| Round 4 | $+1.5 \pm 8.3$ | $-1.7 \pm 7.0$ | $t$ | .470 | +0.43 |

### .3.2 Heart rate SD:SART windows (Round − Training)

**Table 5:** Baseline-corrected mean HR (bpm) during video rest periods between SART rounds. *n*_sham_ = 5, *n*_caff_ = 8.

| Rest period | Sham $\Delta$ | Caff $\Delta$ | Test | $p$ | $d$ |
| --- | --- | --- | --- | --- | --- |
| Pre-Round 2 | $-12.7 \pm 6.6$ | $+2.8 \pm 13.8$ | $t$ | .041* | -1.32 |
| Pre-Round 3 | $-6.2 \pm 13.2$ | $+0.3 \pm 11.4$ | $t$ | .365 | -0.54 |
| Pre-Round 4 | $-10.7 \pm 18.2$ | $-0.8 \pm 8.4$ | $t$ | .204 | -0.77 |
| Post-Round 4 | $-6.6 \pm 7.2$ | $+1.1 \pm 17.5$ | MW | .354 | -0.52 |

### .3.3 Heart rate:rest periods (Epoch − Training)

**Table 6:** Baseline-corrected HR SD (bpm) during video rest periods between SART rounds. *n*_sham_ = 5, *n*_caff_ = 8.

| Rest period | Sham $\Delta$ | Caff $\Delta$ | Test | $p$ | $d$ |
| --- | --- | --- | --- | --- | --- |
| Pre-Round 2 | $+1.3 \pm 6.3$ | $-6.1 \pm 5.8$ | $t$ | .054<sup>†</sup> | +1.23 |
| Pre-Round 3 | $+5.7 \pm 10.8$ | $-6.2 \pm 6.0$ | $t$ | .025* | +1.48 |
| Pre-Round 4 | $+0.4 \pm 15.4$ | $-3.3 \pm 5.7$ | MW | 1.00 | +0.36 |
| Post-Round 4 | $+8.3 \pm 10.7$ | $-2.2 \pm 14.8$ | MW | .065<sup>†</sup> | +0.78 |

### .3.4 Heart rate SD:rest periods (Epoch − Training)

**Table 7:** Baseline-corrected EDA and temperature during SART windows. *n*_sham_ = 7, *n*_caff_ = 9.

| Rnd | Metric | Sham $\Delta$ | Caff $\Delta$ | Test | $p$ | $d$ |
| --- | --- | --- | --- | --- | --- | --- |
| 1 | EDA tonic ($\mu\text{S}$) | $+0.05 \pm 0.11$ | $-1.03 \pm 3.43$ | MW | .299 | +0.42 |
| 1 | SCR rate (/min) | $-18.6 \pm 27.1$ | $+23.0 \pm 35.2$ | $t$ | .022* | -1.30 |
| 1 | Temp ($^{\circ}\text{C}$) | $+1.08 \pm 1.60$ | $+1.21 \pm 1.11$ | $t$ | .854 | -0.09 |
| 1 | Therm ($^{\circ}\text{C}$) | $+1.06 \pm 1.76$ | $+1.30 \pm 0.96$ | $t$ | .737 | -0.17 |
| 2 | EDA tonic ($\mu\text{S}$) | $+0.05 \pm 0.15$ | $-0.44 \pm 4.06$ | MW | .470 | +0.16 |
| 2 | SCR rate (/min) | $-18.0 \pm 29.7$ | $+12.4 \pm 41.3$ | $t$ | .123 | -0.83 |
| 2 | Temp ($^{\circ}\text{C}$) | $+0.89 \pm 1.75$ | $+1.25 \pm 1.31$ | $t$ | .649 | -0.23 |
| 2 | Therm ($^{\circ}\text{C}$) | $+0.81 \pm 1.84$ | $+1.29 \pm 1.14$ | $t$ | .533 | -0.32 |
| 3 | EDA tonic ($\mu\text{S}$) | $+0.04 \pm 0.18$ | $+3.45 \pm 14.48$ | MW | .758 | -0.31 |
| 3 | SCR rate (/min) | $-12.3 \pm 27.0$ | $+9.7 \pm 43.4$ | $t$ | .262 | -0.59 |
| 3 | Temp ($^{\circ}\text{C}$) | $+1.00 \pm 2.05$ | $+1.25 \pm 1.51$ | $t$ | .788 | -0.14 |
| 3 | Therm ($^{\circ}\text{C}$) | $+0.63 \pm 2.08$ | $+1.14 \pm 1.37$ | $t$ | .569 | -0.29 |
| 4 | EDA tonic ($\mu\text{S}$) | $+0.04 \pm 0.19$ | $-1.15 \pm 3.37$ | MW | .837 | +0.47 |
| 4 | SCR rate (/min) | $+0.4 \pm 19.5$ | $+12.7 \pm 38.6$ | MW | 1.00 | -0.39 |
| 4 | Temp ($^{\circ}\text{C}$) | $+0.84 \pm 1.90$ | $+1.00 \pm 1.51$ | $t$ | .850 | -0.10 |
| 4 | Therm ($^{\circ}\text{C}$) | $+0.46 \pm 1.97$ | $+0.93 \pm 1.41$ | $t$ | .584 | -0.28 |

### .3.5 EDA and temperature:SART windows (Round − Training)

**Table 8:** Baseline-corrected EDA and temperature during video rest periods. *n*_sham_ = 7, *n*_caff_ = 9.

| Rest period | Metric | Sham $\Delta$ | Caff $\Delta$ | Test | $p$ | $d$ |
| --- | --- | --- | --- | --- | --- | --- |
| Pre-Rnd 2 | EDA tonic ($\mu\text{S}$) | $+0.03 \pm 0.04$ | $+4.92 \pm 14.92$ | MW | .681 | -0.43 |
| Pre-Rnd 2 | SCR rate (/min) | $+1.8 \pm 14.6$ | $-2.7 \pm 14.3$ | $t$ | .556 | +0.30 |
| Pre-Rnd 2 | Temp ($^{\circ}\text{C}$) | $-0.13 \pm 0.20$ | $+0.17 \pm 0.49$ | $t$ | .156 | -0.76 |
| Pre-Rnd 2 | Therm ($^{\circ}\text{C}$) | $-0.26 \pm 0.34$ | $+0.10 \pm 0.38$ | $t$ | .068<sup>†</sup> | -0.99 |
| Pre-Rnd 3 | EDA tonic ($\mu\text{S}$) | $+0.04 \pm 0.07$ | $+0.36 \pm 0.99$ | MW | .606 | -0.42 |
| Pre-Rnd 3 | SCR rate (/min) | $+9.6 \pm 12.2$ | $-6.8 \pm 21.5$ | MW | .091<sup>†</sup> | +0.91 |
| Pre-Rnd 3 | Temp ($^{\circ}\text{C}$) | $-0.09 \pm 0.51$ | $+0.27 \pm 0.83$ | $t$ | .322 | -0.52 |
| Pre-Rnd 3 | Therm ($^{\circ}\text{C}$) | $-0.35 \pm 0.69$ | $+0.05 \pm 0.76$ | $t$ | .302 | -0.54 |
| Pre-Rnd 4 | EDA tonic ($\mu\text{S}$) | $+0.04 \pm 0.09$ | $-0.12 \pm 0.48$ | MW | .918 | +0.43 |
| Pre-Rnd 4 | SCR rate (/min) | $+2.6 \pm 12.2$ | $-4.9 \pm 18.8$ | $t$ | .379 | +0.46 |
| Pre-Rnd 4 | Temp ($^{\circ}\text{C}$) | $-0.12 \pm 0.80$ | $+0.02 \pm 0.81$ | $t$ | .739 | -0.17 |
| Pre-Rnd 4 | Therm ($^{\circ}\text{C}$) | $-0.51 \pm 0.74$ | $-0.21 \pm 0.75$ | $t$ | .426 | -0.41 |
| Post-Rnd 4 | EDA tonic ($\mu\text{S}$) | $+0.03 \pm 0.10$ | $-0.15 \pm 0.54$ | MW | .918 | +0.44 |
| Post-Rnd 4 | SCR rate (/min) | $+3.3 \pm 7.5$ | $-1.6 \pm 19.1$ | $t$ | .536 | +0.32 |
| Post-Rnd 4 | Temp ($^{\circ}\text{C}$) | $-0.32 \pm 0.66$ | $-0.23 \pm 0.82$ | $t$ | .831 | -0.11 |
| Post-Rnd 4 | Therm ($^{\circ}\text{C}$) | $-0.77 \pm 0.56$ | $-0.42 \pm 0.77$ | $t$ | .327 | -0.51 |

### .3.6 EDA and temperature:rest periods (Epoch − Baseline)

## Footnotes

1 The study was pre-registered (Appendix **??**).

## References

[1] Judith Amores and Pattie Maes. 2017. Essence: Olfactory Interfaces for Unconscious Influence of Mood and Cognitive Performance. In Proceedings of the 2017 CHI Conference on Human Factors in Computing Systems. ACM, 28–34. doi:10.1145/3025453.3026004

[2] Anonymous. 2026. Pre-registration: Wearable sonophoresis for transdermal caffeine delivery. AsPredicted #279566. Filed after data collection but before any statistical analysis. https://aspredicted.org/79c2p9.pdf.

[3] Ruben T. Azevedo, Nell Bennett, Andreas Bilicki, Jack Hooper, Fotini Markopoulou, and Manos Tsakiris. 2017. The Calming Effect of a New Wearable Device During the Anticipation of Public Speech. Scientific Reports 7 (2017), 2285. doi:10.1038/s41598-017-02274-2

[4] Alain Boucaud, Marie Ange Garrigue, Laurent Machet, Loıc Vaillant, and Frédéric Patat. 2002. Effect of sonication parameters on transdermal delivery of insulin to hairless rats. Journal of controlled release 81, 1-2 (2002), 113–119.

[5] Arnaud Boucaud, Laurent Machet, Bertrand Arbeille, Marie Christine Machet, Monique Sournac, Alain Mavon, Frédéric Patat, and Loïc Vaillant. 2001. In Vitro Study of Low-Frequency Ultrasound-Enhanced Transdermal Transport of Fentanyl and Caffeine Across Human and Hairless Rat Skin. International Journal of Pharmaceutics 228, 1–2 (2001), 69–77. doi:10.1016/S0378-5173(01)00820-1

[6] Jas Brooks and Pedro Lopes. 2024. Augmented Breathing via Thermal Feedback in the Nose. In Proceedings of the 37th Annual ACM Symposium on User Interface Software and Technology (UIST ‘24). doi:10.1145/3654777.3676364

[7] Jas Brooks, Pedro Lopes, Judith Amores, Emanuela Maggioni, Haruka Matsukura, Marianna Obrist, Roshan Lalintha Peiris, and Nimesha Ranasinghe. 2021. Smell, Taste, and Temperature Interfaces. In Extended Abstracts of the 2021 CHI Conference on Human Factors in Computing Systems. ACM, 1–6. doi:10.1145/3411763.3441317

[8] Jas Brooks, Steven Nagels, and Pedro Lopes. 2020. Trigeminal-based Temperature Illusions. In Proceedings of the 2020 CHI Conference on Human Factors in Computing Systems (CHI ‘20). 1–12. doi:10.1145/3313831.3376806

[9] Jean Costa, François Guimbretière, Malte F. Jung, and Tanzeem Choudhury. 2019. BoostMeUp: Improving Cognitive Performance in the Moment by Unobtrusively Regulating Emotions with a Smartwatch. Proceedings of the ACM on Interactive, Mobile, Wearable and Ubiquitous Technologies 3, 2 (2019), 1–23. doi:10.1145/3328911

[10] Duramobi. 2024. Duramobi Mini Portable Speaker. https://www.duramobi.com Compact speaker module used as ultrasound transducer.

[11] EmotiBit. 2023. EmotiBit: Open-source wearable for physiological sensing. https://www.emotibit.com/ Accessed: 2025.

[12] Espressif Systems. 2023. ESP32-WROOM-32: Wi-Fi and Bluetooth Module. https://www.espressif.com/en/products/modules/esp32 Dual-core microcontroller with integrated Bluetooth Low Energy.

[13] Stephen H. Fairclough. 2009. Fundamentals of Physiological Computing. Interacting with Computers 21, 1–2 (2009), 133–145. doi:10.1016/j.intcom.2008.10.011

[14] Bertil B. Fredholm, Karl Bättig, Jessica Holmén, Astrid Nehlig, and Edwin E. Zvartau. 1999. Actions of caffeine in the brain with special reference to factors that contribute to its widespread use. Pharmacological Reviews 51, 1 (1999), 83–133.

[15] Asma Ghandeharioun and Rosalind W. Picard. 2017. BrightBeat: Effortlessly Influencing Breathing for Cultivating Calmness and Focus. In Proceedings of the 2017 CHI Conference Extended Abstracts on Human Factors in Computing Systems. ACM, 1624–1631. doi:10.1145/3027063.3053164

[16] Kayla J. Heffernan, Frank Vetere, and Shanton Chang. 2016. Insertables: I’ve Got It Under My Skin. Interactions 23, 1 (2016), 52–56. doi:10.1145/2843590

[17] Kristin E. Heron and Joshua M. Smyth. 2010. Ecological Momentary Interventions: Incorporating Mobile Technology into Psychosocial and Health Behaviour Treatments. British Journal of Health Psychology 15, 1 (2010), 1–39.

[18] Interaxon Inc. 2023. Muse S: The brain sensing headband. https://choosemuse.com/ Accessed: 2025.

[19] Abhinandan Jain, Kyung Yun Choi, Hiroshi Ishii, and Pattie Maes. 2023. Modulating Interoceptive Signals for Influencing the Conscious Experience. In Extended Abstracts of the 2023 CHI Conference on Human Factors in Computing Systems. 1–7. doi:10.1145/3544549.3585791

[20] Abhinandan Jain, Adam Haar Horowitz, Felix Schoeller, Sang-won Leigh, Pattie Maes, and Misha Sra. 2020. Designing Interactions Beyond Conscious Control: A New Model for Wearable Interfaces. Proceedings of the ACM on Interactive, Mobile, Wearable and Ubiquitous Technologies 4, 3, Article 108 (2020), 23 pages. doi:10.1145/3411829

[21] Predrag Klasnja, Eric B. Hekler, Saul Shiffman, Audrey Borber, Daniel Unber, Ricardo Henao, and Susan A. Murphy. 2015. Microrandomized Trials: An Experimental Design for Developing Just-in-Time Adaptive Interventions. Health Psychology 34, S (2015), 1220–1228.

[22] Christian Kothe, Seyed Yahya Shirazi, Tristan Stenner, David Medine, Chadwick Boulay, Matthew I. Grivich, Fiorenzo Artoni, Tim Mullen, Arnaud Delorme, and Scott Makeig. 2025. The Lab Streaming Layer for Synchronized Multimodal Recording. Imaging Neuroscience 3 (2025), IMAG.a.136. doi:10.1162/IMAG.a.136

[23] Richard B. Kreider, Douglas S. Kalman, Jose Antonio, Tim N. Ziegenfuss, Robert Wildman, Rick Collins, Darren G. Candow, Susan M. Kleiner, Anthony L. Almada, and Hector L. Lopez. 2017. International Society of Sports Nutrition Position Stand: Safety and Efficacy of Creatine Supplementation in Exercise, Sport, and Medicine. Journal of the International Society of Sports Nutrition 14 (2017), 18. doi:10.1186/s12970-017-0173-z

[24] Lab Alley. 2024. Caffeine USP Grade Powder. https://www.laballey.com/products/caffeine-usp United States Pharmacopeia certified.

[25] Pedro Lopes. 2025. What if the “I” in HCI Stands for Integration?. In Adjunct Proceedings of the 38th Annual ACM Symposium on User Interface Software and Technology (UIST Adjunct ‘25). doi:10.1145/3708359.3733498

[26] Pedro Lopes and Patrick Baudisch. 2013. Muscle-Propelled Force Feedback: Bringing Force Feedback to Mobile Devices. In Proceedings of the SIGCHI Conference on Human Factors in Computing Systems. 2577–2580. doi:10.1145/2470654.2481355

[27] Pedro Lopes, Alexandra Ion, and Patrick Baudisch. 2015. Impacto: Simulating Physical Impact by Combining Tactile Stimulation with Electrical Muscle Stimulation. In Proceedings of the 28th Annual ACM Symposium on User Interface Software & Technology. 11–19. doi:10.1145/2807442.2807443

[28] Jasmine Lu, Ziwei Liu, Jas Brooks, and Pedro Lopes. 2021. Chemical Haptics: Rendering Haptic Sensations via Topical Stimulants. In Proceedings of the 34th Annual ACM Symposium on User Interface Software and Technology (UIST ‘21). ACM, 239–257. doi:10.1145/3472749.3474747

[29] Jasmine Lu, Ziwei Liu, and Pedro Lopes. 2023. Taste Retargeting via Chemical Taste Modulators. In Proceedings of the 36th Annual ACM Symposium on User Interface Software and Technology (UIST ‘23). doi:10.1145/3586183.3606764

[30] Lin Luo and Majella E. Lane. 2015. Topical and Transdermal Delivery of Caffeine. International Journal of Pharmaceutics 490, 1–2 (2015), 155–164. doi:10.1016/j.ijpharm.2015.05.050

[31] Pattie Maes. 1994. Agents that Reduce Work and Information Overload. Commun. ACM 37, 7 (1994), 30–40. doi:10.1145/176789.176792

[32] D. Marathe, Vasudeva Sampriya Bhuvanashree, C. H. Mehta, T. Ashwini, and U. Y. Nayak. 2024. Low-Frequency Sonophoresis: A Promising Strategy for Enhanced Transdermal Delivery. Advances in Pharmacological and Pharmaceutical Sciences 2024, 1 (2024). doi:10.1155/2024/1247450

[33] Tom M. McLellan, John A. Caldwell, and Harris R. Lieberman. 2016. A review of caffeine’s effects on cognitive, physical and occupational performance. Neuroscience & Biobehavioral Reviews 71 (2016), 294–312. doi:10.1016/j.neubiorev.2016.09.001

[34] Douglas Miller and Robert Niichel. 2021. Advancing high tech drug delivery systems for the treatment of Crohn’s disease. Inflammatory Bowel Diseases 27 (2021), S2.

[35] Varun Mishra, Florian Künzler, Jan-Niklas Kramer, Elgar Fleisch, Tobias Kowatsch, and David Kotz. 2021. Detecting Receptivity for mHealth Interventions in the Natural Environment. In Proceedings of the ACM on Interactive, Mobile, Wearable and Ubiquitous Technologies, Vol. 5. 1–24.

[36] Samir Mitragotri, Daniel Blankschtein, and Robert Langer. 1995. Ultrasound-Mediated Transdermal Protein Delivery. Science 269, 5225 (1995), 850–853. doi:10.1126/science.7638603

[37] Samir Mitragotri and Joseph Kost. 2000. Low-Frequency Sonophoresis: A Noninvasive Method of Drug Delivery and Diagnostics. Biotechnology Progress 16, 3 (2000), 488–492. doi:10.1021/bp000024+

[38] Sehreen Moorat, Ahsan Ahmad Ursani, Aftab Ahmed Memon, and Muhammad Aamir Panhwar. 2024. Optimizing Transdermal Insulin Delivery: A Simulation Study on the Efficacy of Sonophoretic Transducer Arrays at Low Voltages. IEEE Access 12 (2024), 115055–115063.

[39] Florian Floyd Mueller, Pedro Lopes, Paul Strohmeier, Wendy Ju, Caitlyn Seim, Martin Weigel, Suranga Nanayakkara, Marianna Obrist, Zhuying Li, Joseph Delfa, Jun Nishida, Elizabeth M. Gerber, Dag Svanaes, Jonathan Grudin, Stefan Greuter, Kai Kunze, Thomas Erickson, Steven Greenspan, Masahiko Inami, Joe Marshall, Harald Reiterer, Katrin Wolf, Jochen Meyer, Thecla Schiphorst, Dakuo Wang, and Pattie Maes. 2020. Next Steps for Human-Computer Integration. In Proceedings of the 2020 CHI Conference on Human Factors in Computing Systems (CHI ‘20). 1–15. doi:10.1145/3313831.3376242

[40] Inbal Nahum-Shani, Shawna N. Smith, Bonnie J. Spring, Linda M. Collins, Katie Witkiewitz, Ambuj Tewari, and Susan A. Murphy. 2018. Just-in-Time Adaptive Interventions (JITAIs) in Mobile Health: Key Components and Design Principles for Ongoing Health Behavior Support. Annals of Behavioral Medicine 52, 6 (2018), 446–462.

[41] Takuji Narumi, Shinya Nishizaka, Takashi Kajinami, Tomohiro Tanikawa, and Michitaka Hirose. 2011. Augmented Reality Flavors: Gustatory Display Based on Edible Marker and Cross-Modal Interaction. In Proceedings of the SIGCHI Conference on Human Factors in Computing Systems (CHI ‘11). 93–102. doi:10.1145/1978942.1978957

[42] Revital Nimri, Judith Nir, and Moshe Phillip. 2020. Insulin pump therapy. American journal of therapeutics 27, 1 (2020), e30–e41.

[43] Dag Nyholm. 2006. Pharmacokinetic optimisation in the treatment of Parkinson’s disease: an update. Clinical pharmacokinetics 45, 2 (2006), 109–136.

[44] Jongsung Park, Hyunjin Lee, Gyeong-Sik Lim, Nari Kim, Donghee Kim, and Young-Chang Kim. 2019. Enhanced Transdermal Drug Delivery by Sonophoresis and Simultaneous Application of Sonophoresis and Iontophoresis. AAPS PharmSciTech 20, 3 (2019), 96. doi:10.1208/s12249-019-1309-z

[45] Baris E. Polat, Douglas Hart, Robert Langer, and Daniel Blankschtein. 2011. Ultrasound-Mediated Transdermal Drug Delivery: Mechanisms, Scope, and Emerging Trends. Journal of Controlled Release 152, 3 (2011), 330–348. doi:10.1016/j.jconrel.2011.01.006

[46] Nada Pop-Jordanova and Jordan Pop-Jordanov. 2005. Spectrum-weighted EEG frequency (“brain-rate”) as a quantitative indicator of mental arousal. Prilozi 26, 2 (2005), 35–42.

[47] Ivan Poupyrev, Nan-Wei Gong, Shiho Fukuhara, Mustafa Emre Karagozler, Carsten Schwesig, and Karen E. Robinson. 2016. Project Jacquard: Interactive Digital Textiles at Scale. In Proceedings of the 2016 CHI Conference on Human Factors in Computing Systems. ACM, 4216–4227. doi:10.1145/2858036.2858176

[48] Mark R. Prausnitz. 2004. Microneedles for Transdermal Drug Delivery. Advanced Drug Delivery Reviews 56, 5 (2004), 581–587. doi:10.1016/j.addr.2003.10.023

[49] Mark R. Prausnitz and Robert Langer. 2008. Transdermal Drug Delivery. Nature Biotechnology 26, 11 (2008), 1261–1268. doi:10.1038/nbt.1504

[50] Ian H. Robertson, Tom Manly, Jackie Andrade, Alan Baddeley, and Jenny Yiend. 1997. ‘Oops!’: Performance correlates of everyday attentional failures in traumatic brain injured and normal subjects. Neuropsychologia 35, 6 (1997), 747–758. doi:10.1016/S0028-3932(97)00015-8

[51] Paul Seli, James Allan Cheyne, Keith R. Barton, and Daniel Smilek. 2013. Consistency of sustained attention across modalities: Comparing visual and auditory versions of the SART. Canadian Journal of Experimental Psychology 67, 1 (2013), 44–52. doi:10.1037/a0031007

[52] Andrew Smith. 2002. Effects of caffeine on human behavior. Food and Chemical Toxicology 40, 9 (2002), 1243–1255. doi:10.1016/S0278-6915(02)00096-0

[53] Joshua M. Smyth and Kristin E. Heron. 2016. Is Providing Mobile Interventions “Just-in-Time” Helpful? An Experimental Proof of Concept Study of Just-in-Time Intervention for Stress Management. IEEE Wireless Health (2016).

[54] Misha Sra, Abhinandan Jain, and Pattie Maes. 2019. Adding Proprioceptive Feedback to Virtual Reality Experiences Using Galvanic Vestibular Stimulation. In Proceedings of the 2019 CHI Conference on Human Factors in Computing Systems (CHI ‘19). 1–14. doi:10.1145/3290605.3300905

[55] Cordula Stillhart, Kristína Vučićević, Patrick Augustijns, Abdul W. Basit, Hannah Batchelor, Talia R. Flanagan, Isabelle Gesquiere, Rick Greupink, Daniel Keszthelyi, Mirko Koskinen, et al. 2021. Impact of Gastrointestinal Tract Variability on Oral Drug Absorption and Pharmacokinetics: An UNGAP Review. European Journal of Pharmaceutical Sciences 162 (2021), 105812. doi:10.1016/j.ejps.2021.105812

[56] Swartz Center for Computational Neuroscience. 2024. Lab Recorder: Recording application for LSL streams. https://github.com/labstreaminglayer/App-LabRecorder Part of the Lab Streaming Layer (LSL) ecosystem for synchronized multi-modal data collection.

[57] Liang-Chia Tai, Wei Gao, Mallika Chao, Merissa Bariya, Quynh Phuong Nez, Ziba Shahpar, Hnin Y. Y. Nyein, Hoseok Park, Jia Sun, Yongkuk Jung, Eric Wu, Hossain M. Fahad, Der-Hsien Lien, Hiroki Ota, Geunjae Cho, and Ali Javey. 2018. Methylxanthine drug monitoring with wearable sweat sensors. Advanced Materials 30, 23 (2018), 1707442. doi:10.1002/adma.201707442

[58] Ahmet Tezel and Samir Mitragotri. 2003. Interactions of Inertial Cavitation Bubbles with Stratum Corneum Lipid Bilayers during Low-Frequency Sonophoresis. Biophysical Journal 85, 6 (2003), 3502–3512. doi:10.1016/S0006-3495(03)74770-5

[59] Ahmet Tezel, Ashley Sens, Jose Tuchscherer, and Samir Mitragotri. 2001. Frequency Dependence of Sonophoresis. Pharmaceutical Research 18, 12 (2001), 1694–1700. doi:10.1023/A:1013366328457

[60] Thorne Research. 2024. Creatine Monohydrate Powder. https://www.thorne.com/products/dp/creatine NSF Certified for Sport.

[61] U.S. Department of Agriculture. 2024. FoodData Central: Coffee, brewed. https://fdc.nal.usda.gov/food-details/171890/nutrients. Accessed: 2026-03-31. Reports 96 mg caffeine per 8 fl oz (240 mL) serving.

[62] Francisco G. Vital-Lopez, Tracy J. Doty, Ian Anlap, William D. S. Killgore, and Jaques Reifman. 2023. 2B-Alert App 2.0: Personalized Caffeine Recommendations for Optimal Alertness. SLEEP 46, 7 (2023), zsad080. doi:10.1093/sleep/zsad080

[63] Francisco G. Vital-Lopez, Sridhar Ramakrishnan, Tracy Jill Doty, Thomas J. Balkin, and Jaques Reifman. 2018. Caffeine Dosing Strategies to Optimize Alertness During Sleep Loss. Journal of Sleep Research 27, 5 (2018), e12711. doi:10.1111/jsr.12711

[64] Lei Wang, Chen Ma, Xueyang Feng, Zeyu Zhang, Hao Yang, Jingsen Zhang, Zhiyuan Chen, Jiakai Tang, Xu Chen, Yankai Lin, Wayne Xin Zhao, Zhewei Wei, and Ji-Rong Wen. 2024. A Survey on Large Language Model based Autonomous Agents. Frontiers of Computer Science 18, 6 (2024), 186345. doi:10.1007/s11704-024-40231-1

[65] Yihang Wang, Tianyu Li, Yang Li, Ruixing Yang, and Guorui Zhang. 2024. Digital Automation of Transdermal Drug Delivery with High Spatiotemporal Resolution. Nature Communications 15 (2024), 640. doi:10.1038/s41467-023-44532-0

[66] Graham Wilson, Martin Halvey, Stephen A. Brewster, and Stephen A. Hughes. 2011. Some Like It Hot: Thermal Feedback for Mobile Devices. In Proceedings of the SIGCHI Conference on Human Factors in Computing Systems (CHI ‘11). 2555–2564. doi:10.1145/1978942.1979316

[67] Chuanqi C. Yu, Anh Vu, Sarvesh Dhiraj, Duc Thinh Le, and Canan Dagdeviren. 2023. A Conformable Ultrasound Patch for Cavitation-Enhanced Transdermal Cosmeceutical Delivery. Advanced Materials 35, 23 (2023), 2300066. doi:10.1002/adma.2300066

